# Implementation of a multi-modal training program for the management of comorbid mental disorders in drug and alcohol settings: Pathways to Comorbidity Care (PCC)

**DOI:** 10.1101/2021.03.18.21253927

**Authors:** Eva Louie, Kirsten C. Morley, Vicki Giannopoulos, Gabriela Uribe, Katie Wood, Christina Marel, Katherine L Mills, Maree Teesson, Michael Edwards, Steven Childs, David Rogers, Adrian Dunlop, Andrew Baillie, Paul S. Haber

## Abstract

**Background:** Clinical guidelines recommend evidence-based treatments for comorbid mental and substance use disorders but these are not reliably translated into practice. We aimed to evaluate the impact of the Pathways to Comorbidity Care (PCC) training program for alcohol and other drug (AOD) clinicians to improve the management of comorbidity and to identify barriers and facilitators of implementation according to the Consolidated Framework for Implementation Research (CFIR).

**Methods:** A controlled before-and-after study using PCC training was conducted across 6 matched sites in Australia including 35 clinicians. Controls received standard workplace training. PCC training included seminar presentations, workshops conducted by local ‘clinical champions’, individual clinical supervision, and access to an online information portal. A mixed methods approach examined i) identification (screening, assessment) and treatment (treatment, referral) of comorbidity in practice (N = 10 clinical files per clinician), ii) self-efficacy, knowledge and attitudes of clinicians, iii) barriers and facilitators of implementation.

**Results:** Significant improvements were observed in the PCC group but not the control sites with regards to the rate of clinical files showing identification of comorbidity (+50% v −12% change from baseline respectively; *X*^2^ (1, *N* = 340) = 35.29, p = .01) with only a trend for improvements in the rate of files demonstrating treatment of comorbidity (*X*^2^ (1, *N* = 340) = 10.45, p = .06). There were significant improvements in the PCC relative to the control group for clinician self-efficacy (F (1,33) = 6.40, p = .02) and knowledge and attitudes of comorbidity monitoring (F (1,33) = 8.745, p = .01). Barriers included inner setting (e.g. allocated time for learning) and characteristics of individuals (e.g. resistance). Facilitators included intervention characteristics (e.g. credible sources), inner setting (e.g. leadership) and outer setting domains (e.g. patient needs). Clinical champions were identified as an important component of the implementation process.

**Conclusions:** The PCC training package effectively improved identification of comorbidity, self-efficacy and attitudes towards screening and monitoring of comorbidity. Specific barriers included provision of allocated time for learning. Specific facilitators included provision of a credible clinical supervisor, strong leadership engagement and an active clinical champion.

## Introduction

There is a high degree of comorbid mental disorders in alcohol and other drug (AOD) treatment settings. Up to 90% of people accessing substance use treatment also experience comorbid mental health problems ^1^. Comorbid mental disorders and AOD use is associated with greater symptom severity, reduced quality of life and increased reliance on treatment services compared to those with AOD alone ^2,3^, and pose a significant challenge for AOD services. The non-integrated nature of many mental health and substance use services also presents challenges for those intending to access appropriate services ^4^. Consequently, many people do not receive effective interventions and experience poor clinical outcomes ^2,3^.

One evidence-based model for managing comorbidity is integrated care. Integrated care aims to provide coordinated, efficient and effective care that responds to all of the needs of the client. An integrated model requires identification and assessment of both the AOD use and the mental health condition, along with a comprehensive management plan for treating both problems ^5^. Offering this integrated care within one service overcomes problems associated with non-integrated treatment, including physical barriers (such as services located separately) and non-cohesive treatment plans. Although integrated care may not always be appropriate, this approach is often considered the preferred model of care and has been widely recommended in clinical guidelines ^5^.

Specific psychological interventions employed by substance use clinicians in an integrated care approach include specific cognitive behavioural therapy (CBT) and motivational interviewing (MI) techniques ^6^. There is evidence for the efficacy of specific CBT-based integrated treatments for comorbid depression and substance use ^7,8^, anxiety ^9^, anxiety and depression ^10^ and post-traumatic stress disorder (PTSD) ^9,11,12^. While extensive evidence for the efficacy of treatments for psychosis with comorbid substance use disorder is lacking ^13,14^, integrated psychological and pharmacological treatments are generally supported ^15,16^.

Implementing integrated care on a large scale in public health treatment settings is a complex process, given the broad range of clinical training and experience, health service leadership, organisational dynamics and health systems ^17^. Evidence for the effectiveness of integrated treatment for comorbid substance use and mental disorders has often been derived from randomised controlled trials utilising independent and highly trained clinicians employed specifically for these trials ^10,18,19^ rather than clinicians in practice. Clinicians practicing in AOD treatment settings may not have undergone training in the identification and treatment of mental health disorders and may view these disorders as outside the scope of their role, while some managers may view that the management of mental disorders is beyond the capacity of the service. Facilitating the implementation of integrated care, when it is appropriate, thus requires building capacity in these settings.

The multi-modal Pathways to Comorbidity Care (PCC) training package was thus developed to target potential barriers to delivering integrated care for comorbid substance use and mental disorders in AOD settings. These included improving knowledge, attitudes and confidence of AOD clinicians to manage these problems see ^20^. We developed the PCC training to occur via multiple modalities, designed to present didactic material to establish a standard of knowledge (resources, seminars) followed by provision of interactive learning (clinical supervision and clinical champions) to problem solve implementation in these settings. There have been two previous studies evaluating specific training programs for managing patients with comorbid substance use and mental disorders with mixed results ^21,22^. More broadly in AOD settings, multi-level strategies rather than single level strategies, such as those that focus only on the provider, have previously been found to be preferable to facilitate integrated care ^23^. Indeed, bridging the gap between evidence and practice requires systematic assessment of the implementation barriers that exist at multiple levels of healthcare delivery including the patient level, the provider level and the organisational level^24^.

To this degree, the Consolidated Framework for Implementation Research (CFIR) ^25^ has been suggested to be suitable model to guide systematic evaluation of multi-level implementation contexts ^26^. The CFIR includes five domains of influence derived from a consolidation of the plethora of terms and concepts generated by implementation researchers: (1) intervention characteristics (e.g. evidence strength and quality, adaptability), (2) outer setting (e.g. patient needs and resources, external policies and incentives), (3) inner setting (e.g. implementation climate, readiness for implementation), (4) individuals involved (e.g. self-efficacy, knowledge and beliefs about the intervention), and (5) the implementation process (e.g. engaging members of the organisation, executing the innovation). No previous studies have systematically evaluated barriers and facilitators of implementation of comorbidity training according to a validated framework, which is key to refining ongoing training programs and future roll out efforts ^27,28^.

This study aimed to i) evaluate the implementation of the PCC training package to improve clinician practice (identification and treatment), confidence (self-efficacy), knowledge and attitudes with regards to comorbid substance use and mental disorders; and ii) identify barriers and facilitators of the PCC program using the CFIR. The CFIR was employed as a guiding framework for determining the specificities of the implementation context, evaluating the implementation and providing a means of assessing the outcome of the implementation.

## Methods

### Study design

This was a controlled, before-and-after study (0-9 months) comparing PCC-training versus control regarding the uptake of comorbidity management. Three PCC and three control sites were matched according to geographical location across six government AOD outpatient and community health services in NSW, Australia (June 2017-2018). Ethical approval for the study was obtained from the Human Ethics Review Committees of the Sydney Local Health District, South Western Sydney Local Health District, Central Coast Local Health District, Hunter New England Research Ethics and Governance Office which covered two participating services, and Mid North Coast Local Health District (X16-0440 & HREC/16/RPAH/624).

### Participants

At each site, all clinicians currently performing an AOD counselling role were invited to participate in the study. A signed buy-in from the managers of each site was obtained including a statement that the organisation has endorsed the use of integrated comorbidity management including support for time and resources for clinicians to participate. Potential clinical champions were identified by managers at each PCC site.

### Study procedures

All participants provided informed consent before taking part in the study. An interview with directors or managers from each of the sites was conducted at baseline. Research assessments were conducted at baseline, 3, 6, and 9 months. Approximately 12 months after baseline, semi-structured interviews were conducted with clinicians at PCC sites.

### Pathways to Comorbidity Care intervention

Participants at PCC sites commenced the training program after completing the baseline assessment. The PCC training program has been described in detail previously ^20^ and thus is outlined only briefly below.

Phase 1 (Months 1-3): This was a 12-week non-intensive period of training whereby participants were given access to the online training portal ^29^ containing various comorbidity resources, the National Comorbidity Guidelines ^5^ and manuals. Within the following month, a one-day face to face seminar was conducted at each of the PCC sites (including webinars about comorbid Substance Use and Depression, Anxiety, Trauma, Psychosis and Bipolar Disorder).

Phase 2 (Months 3-6): This was a 12-week intensive period in which local clinical champions conducted regular group workshops and clinicians received telephone supervision from an experienced senior clinical psychologist ^30^.

Phase 3 (Months 6-9): Participants were provided prompts to revisit the training portal www.pccportal.org.au. Webinars from Phase 1 were also made available on the portal as booster sessions.

### Control sites

Control sites received the standard workplace training as per usual supervision arrangements.

### Assessments

At baseline, participating clinicians completed a questionnaire package including: an adapted version of the Personnel Data Inventory (an index for obtaining demographic and professional information from a pool of clinicians; ^31^); the Evidence-Based Practice Attitudes Scale (EBPAS; ^32^) which measures four aspects of attitude towards evidence based practice (i.e. intuitive appeal, likelihood of adopting if required to, openness to new practices and perceived divergence of practice with evidence-based practice); the Survey of Attitudes to Therapist Manuals which addresses experience with treatment manuals, attitudes towards treatment manuals, and beliefs about the content of treatment manuals ^33^; the Addiction Counseling Self-Efficacy Scale (ACSES: ^34^) which assesses addiction counsellors’ self-efficacy when working with patients including comorbid presentations; the Comorbidity Guidelines Survey (CGS) which were developed for and used in previous evaluations of comorbidity resources and measures clinician knowledge and attitudes regarding comorbidity ^35^ and the Organizational Readiness for Change Assessment Tool (ORCA: ^36^) which measures organisational readiness for implementing practice change in healthcare settings and maps onto the CFIR. The ACSES and CGS were also administered after Phase 1 and 2 for the PCC sites. After phase 3, the final follow-up assessment included AC-SES, CGS and ORCA. Specific questions asking respondents to evaluate Phase 1, 2 and 3 of the PCC Package were included in the questionnaires distributed.

For both PCC and control sites, file audits of clinical notes were conducted at baseline and at follow-up and included clinical notes that were made during the three months prior to each time point (10 files per clinician). A checklist for comorbidity practice (CP) as aligned with ^37^ was used to identify relevant practice themes including screening and monitoring, assessment, treatment and referral which were compressed into two relevant practice themes:

1. Identification (screening and assessment) and 2. Treatment (treatment and referral).

A semi-structured interview was conducted with each participating clinician in the PCC sites at follow-up and evaluated according to the CFIR. The CFIR consolidates the concepts generated by implementation research into five domains of influence: (1) intervention characteristics, (2) outer setting, (3) inner setting, (4) individuals involved, and (5) the implementation process ^38^.

### Outcomes measures

Primary outcomes: Comorbidity practice (CP) assessed using the comorbidity checklist on file audit data as per above), Clinician self-efficacy (as measured by ACSES), knowledge and attitudes (CGS). We also examined predictors of change in self-efficacy: age, professional role, education and training, professional experience, preferred therapeutic modalities, attitudes to therapist manuals (SATM), and attitudes to evidence based practice (EBPAS).

Barriers and facilitator outcomes: intervention characteristics, outer setting, inner setting, characteristics of individuals and the implementation process as per the CFIR (see below).

### Coding

Interviews were transcribed (KW, GU, EL) and coded using thematic analysis to identify perspectives and themes, with the CFIR providing a guiding framework for interpretation.

We developed one codebook before coding the data. In the codebook, we initially included all 39 CFIR constructs and their definitions as codes to capture contextual factors that might influence the implementation of PCC components. These CFIR codes were analytical in that they required the coder to interpret the data and then apply the CFIR code that reflected a potential barrier or facilitator being described. The identification of barriers and facilitators was the main theoretical driver of our study. We applied the CFIR codes to fit the context of the PCC training program by first creating a set of structured and semi-\ structured interview questions that related directly to the PCC intervention and then identifying which of the CFIR domains were addressed. Consequently, certain subdomains of the CFIR were missing (e.g. inner setting characteristics including tension for change and readiness for implementation were not assessed).

Responses were coded by EL using a directed content analysis approach ^39^ in which responses were placed in the most relevant domain. If a response could be coded into more than one domain, EL allocated the most appropriate domain. The coding of the interviews was checked by other team members (KM, KW, GU).

### Analysis

Continuous and categorical baseline variables were examined with Chi square tests and ANOVAs respectively to detect any significant differences in demographic, education and professional experience between control and PCC sites. Two comorbidity practice themes were evaluated for each of Identification and Treatment (present/absent for each item as per the clinician CP checklist) and analysed using McNemar tests for the percentage of total files (N = 340). The outcomes self-efficacy (ACSES) and knowledge and attitude (from the CGS) variables were entered separately into repeated measures ANOVAs comparing PCC versus control. Individual items on the scale of the CGS were analysed separately with a reduced p threshold given that factor analysis and principal components analyses revealed the overall score was limited. Bivariate correlations were conducted to identify and examine the relationship between change in the continuous outcome of self-efficacy (ACSES) and a number of potential baseline characteristics. These variables included *demographic*: age, sex, highest degree; *professional characteristics*: professional role, time in current role, organisational readiness score; *individual characteristics:* Evidence-Based Practice Scores. Any variables associated with primary outcomes with p ≤ 0.10 were placed in the regression model for that outcome. Multiple linear regression analyses were then performed including predictor variables salient in bivariate correlations. All analyses were 2-tailed, with significance level at p < 0.05, apart from the CGS knowledge and attitudes items which were evaluated at a significance level of p < 0.01 as outlined above. Data were analysed using SPSS 24 for Mac OSX.

To analyse the coded data for the barriers and facilitators of the PCC program, we generated code reports from NVIVO software for each transcription for each combination of PCC component and CFIR construct. Within each report, data was organised by CFIR domain (e.g. intervention characteristics). We then developed analytic summaries for each CFIR construct and determined whether the component exerted either a positive (strength) or negative (weakness) on implementation.

## RESULTS

### Sample characteristics

Six AOD services including 3 PCC and 3 control sites participated. Out of 45 clinicians that consented to be involved in the study, 35 participants (N = 20 PCC, N = 15 control) completed the study training (see Figure 1 for flow of participants).

**Figure 1.**
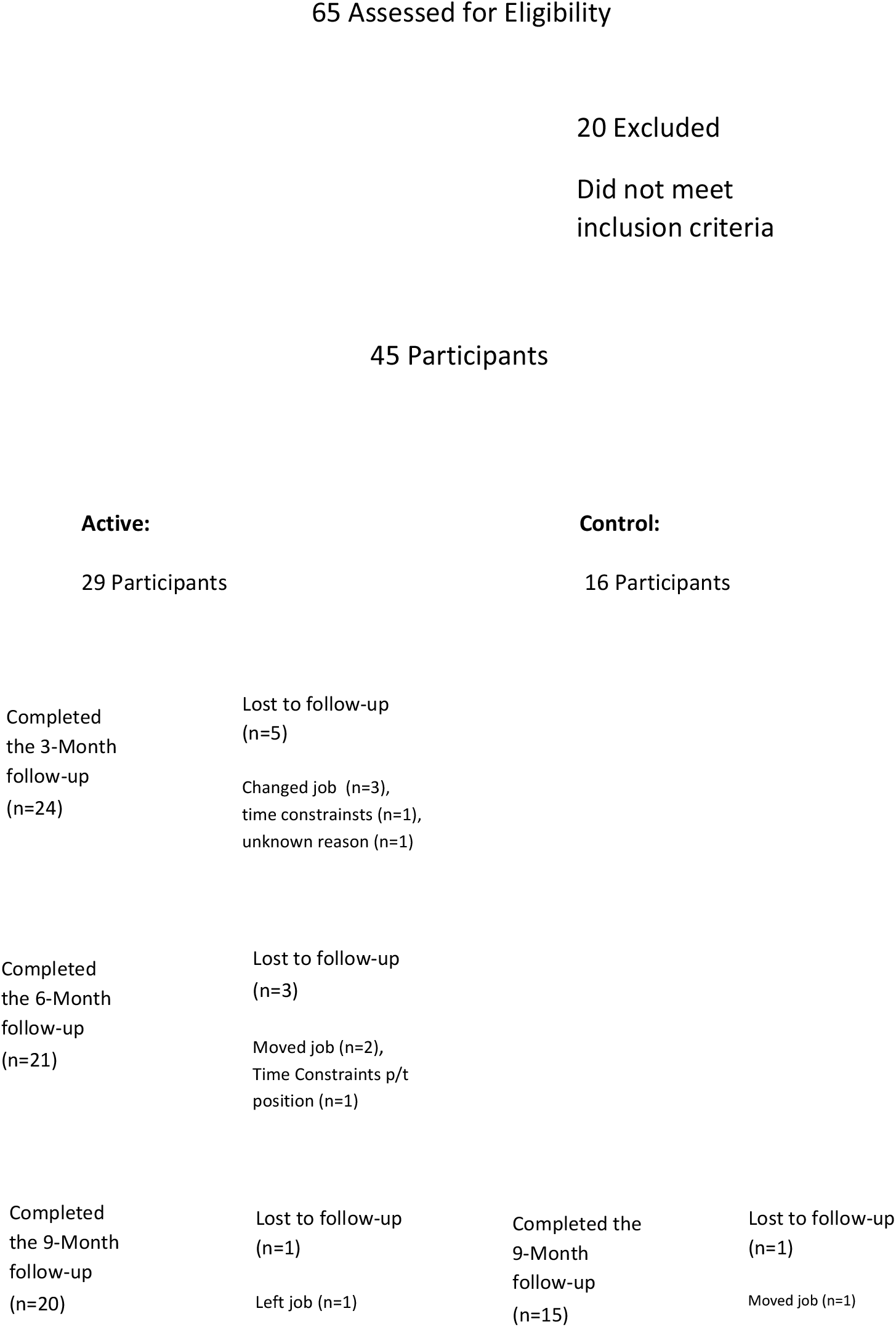
Flow of participants through the Pathways to Comorbidity Care (PCC) study (Active = PCC training at the site, Control = standard training at the site)

Baseline characteristics are displayed in Table 1. The overall mean age was 48.56 (SD ±10.82) years, and 65.7% were female. The majority of participants (62.9%) had completed a university degree, the most common professional role was psychologist (42.9%) and almost all participants (91.4%) had done some form of training in mental health. There were no significant differences between PCC and control groups on measures at baseline (Fs < 1.67) except for part-time staff (*X*^2^ (1, *N* = 35) = 4.23, *p* = .04; PCC = 80% vs control = 47%).

**Table 1.** Baseline characteristics

| **Characteristic** | **PCC**<br>(n = 20) | **Control**<br>(n = 15) |
| --- | --- | --- |
| Mean Age, y | 51.53 +/- 8.14 | 45.20 +/-12.68 |
| Gender, % F | 75 | 53.3 |
| **Education:** |  |  |
| Mean Years since graduating, y | 16.10 +/- 9.13 | 14.60+/-8.77 |
| Highest Degree, % |  |  |
| Bachelor's | 60 | 67 |
| Master's | 30 | 13 |
| Doctorate | 10 | 7 |
| **Professional Role: %** |  |  |
| Psychologist | 45 | 40 |
| Nurse | 10 | 13 |
| Counsellor | 15 | 20 |
| Social Worker | 15 | 20 |
| Case Worker | 10 | 0 |
| **Training: %** |  |  |
| MH training last 12 months | 55 | 47 |
| **Professional Experience: %** |  |  |
| Years of experience in D&A or MH<br>(modal category+) | > 15 years | 5-10 years |
| Years of experience in comorbid<br>D&A and MH (modal category+) | 5-10 years, > 15<br>years | 5-10 years |
| **Practicing nonevidence-**<br>**intervention %** | 20 | 13 |
| **Current Position: %** |  |  |
| Employment status, PT* | 80 | 47 |
| **Measures:** |  |  |
| EBPAS | 59.75 +/- 8.31 | 58.80 ± 6.28 |
| SATM |  |  |
| Negative | 29.75 +/- 7.77 | 28.80 +/- 8.32 |
| Positive | 22.95 +/- 4.59 | 24.20 +/- 3.28 |
| ACSES | 123.15 +/- 18.78 | 124.13 +/- 14.44 |
| ORCA | 76.75 +/- 15.92 | 77.00 +/- 9.37 |

Notes: Data represent mean ± SD unless otherwise noted. \* $p < 0.05$ , significant difference between groups, Chi square. Abbreviations: MH = Mental Health, D&A = Drug and Alcohol, EBPAS = Evidence-Based Practice Attitudes Scale, SATM = the Survey of Attitudes to Therapist Manuals, ACSES = the Addiction Counseling Self-Efficacy Scale (range 32-160), ORCA = the Organizational Readiness for Change Assessment Tool, + = categories of experience included 0-1 year, 1-5 years, 5-10 years, 10-15 years, > 15 years. Nonevidence-based interventions = participant identified currently practicing a therapeutic intervention that is nonevidence-based.

### Retention and compliance

Overall, 78% of the initial sample (N = 45) completed the study. Reasons for drop-out included changes in employment, time constraints and unknown reasons (Figure 1). Of the 35 participants who completed the protocol, 32 (91%) completed all aspects of the training and research tasks.

### Clinician outcomes

#### Clinician comorbidity practice (CP)

Clinical notes were not able to be retrieved for one clinician such that only files of 34 clinicians were reviewed. Baseline and follow-up and changes in CP are depicted in Table 2. Examining the percentage of total clinical files demonstrating Identification (determined via the CP checklist including screening and assessment of symptoms associated with mental disorders) and Treatment (determined via the CP checklist including referral and treatment of mental disorder) revealed significant differences in the PCC group baseline versus follow-up regarding Identification ((*X*^2^ (1, *N* = 340) = 35.29, p = .01, increase) and a near significant trend for Treatment (*X*^2^ (1, *N* = 340) = 10.45, p = .06, increase). There were no significant differences for the control sites regarding either Identification or Treatment from baseline to follow-up (p’s > .24).

**Table 2.** Rate of Identification and Treatment of comorbidity observed in clinical files, before and after receiving the PCC training package or control, per total clinician files.

|  |  | Percentage of total files (N = 340)<br>demonstrating CP |  |
| --- | --- | --- | --- |
|  | Time | PCC sites | Control sites |
| Identification (CP checklist),<br>% (n), | Baseline | 14 (27/190) | 41 (62/150) |
|  | Follow-up | 21 (40/190)* | 36 (54/150) |
| Treatment (CP checklist), %<br>(n), | Baseline | 29 (55/190) | 24 (34/150) |
|  | Follow-up | 34 (64/190) | 28 (42/150) |

Notes: CP = Comorbidity Practice; SD = Standard Deviation; PCC = Pathways to Comorbidity Care. PCC consisted of a one-day seminar, clinical champion run workshops, individual telephone supervision, and online portal containing comorbidity resources. Results represent % = percentage of total clinical files demonstrating CP as defined by Identification (determined via the CP checklist including screening and assessment of symptoms associated with a mental disorder) and Treatment (determined via the CP checklist including referral and treatment of mental disorder). \* $p < 0.05$ , significant increase within group McNemar test (Baseline vs Follow-up).

#### Self-efficacy

Baseline and follow-up ACSES are depicted in Table 3. Repeated measures ANOVA revealed a significant interaction effect of time (baseline to 9-months follow-up) and group (controls versus PCC) in clinician self-efficacy (Wilks’ Lambda = .84, F (1,33) = 6.40, p = .016). Follow-up t-tests of ACSES scores revealed no significant increase from baseline (M=123.15, SD=18.79) to Phase 1 (M=122.60, SD=19.57; t(19=.214, p=.833)) nor Phase 2 (M=129.68, SD=17.13) and 3 (follow-up) (M=133.25, SD=16.32; t(18=-2.077, p=.052)) but a significant increase between Phase 1 (M=122.60, SD=19.57) and 2 (M=129.68, SD=17.13) was revealed (t(18=-2.44, p=.025)). Multiple linear regression of relevant baseline characteristics (age, professional role, attitudes to evidence based practice) revealed that only professional role (psychologist) (β=9.63, p<.01) and amount of experience working with substance use and mental disorders (β=-3.95, p<.05) significantly predicted significant increases in ACSES from baseline (R^2^ = .56, F(3,18)=6.32, p<.01).

**Table 3.** Clinician Addiction Counselor Self-efficacy (ACSES) scores before and after receiving the PCC training package or control.

|  | Time | PCC | Control |
| --- | --- | --- | --- |
| Self-efficacy<br>(ACSES), * | Baseline | 123.15 +/- 18.79 | 124.13 +/- 14.44 |
|  | Follow-up | 133.25 +/- 16.32 | 124.33 +/- 12.17 |

Notes: Scores represent mean +/- SD. ACSES = Addiction Counseling Self-Efficacy Scale (range 32-160); CP = Comorbidity Practice; SD = Standard Deviation; PCC = Pathways to Comorbidity Care. PCC consisted of a one-day seminar, clinical champion run workshops, individual telephone supervision, and online portal containing comorbidity resources; \* $p < 0.05$ , intervention (PCC vs Control) x time (Baseline vs Follow-up) repeated measures ANOVA.

#### Knowledge and attitudes

There were significant increases between baseline and follow-up between PCC relative to controls on the item “Mental health symptoms need to be monitored throughout treatment” (Wilks’ Lambda = .79, F (1,33) = 8.745, p = .01). There were no significant differences between PCC and control on other items using a reduced significance p value of 0.01 (p’s > .03).

Barriers and facilitators according to Consolidated Framework for Implementation Research (CFIR)

#### Relevant theoretical domains

All barriers and facilitators could be identified within the CFIR ^38^. Of the 39 CFIR subdomains, 27 were important in understanding the barriers and facilitators to implementing the PCC. Table 4 lists the CFIR domains and corresponding PCC components and whether that domain and component was an implementation weakness or strength. These are briefly outlined below, and the facilitators are summarised in Figure 2. Agreement between ratings of the CFIR subdomains was 100%, and disagreement was 0% respectively.

**Table 4.**
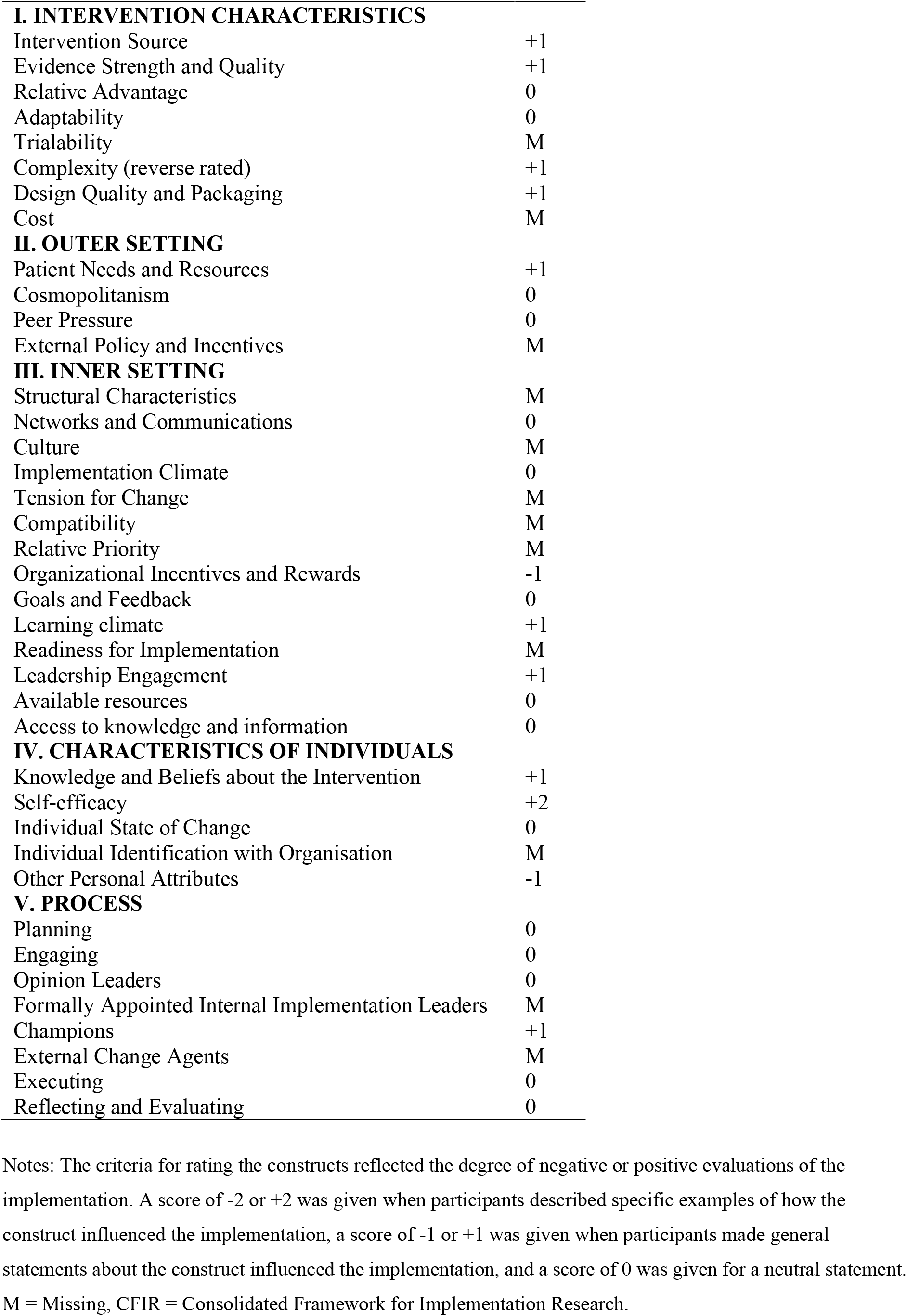
Ratings assigned to CFIR construct

**Figure 2.**
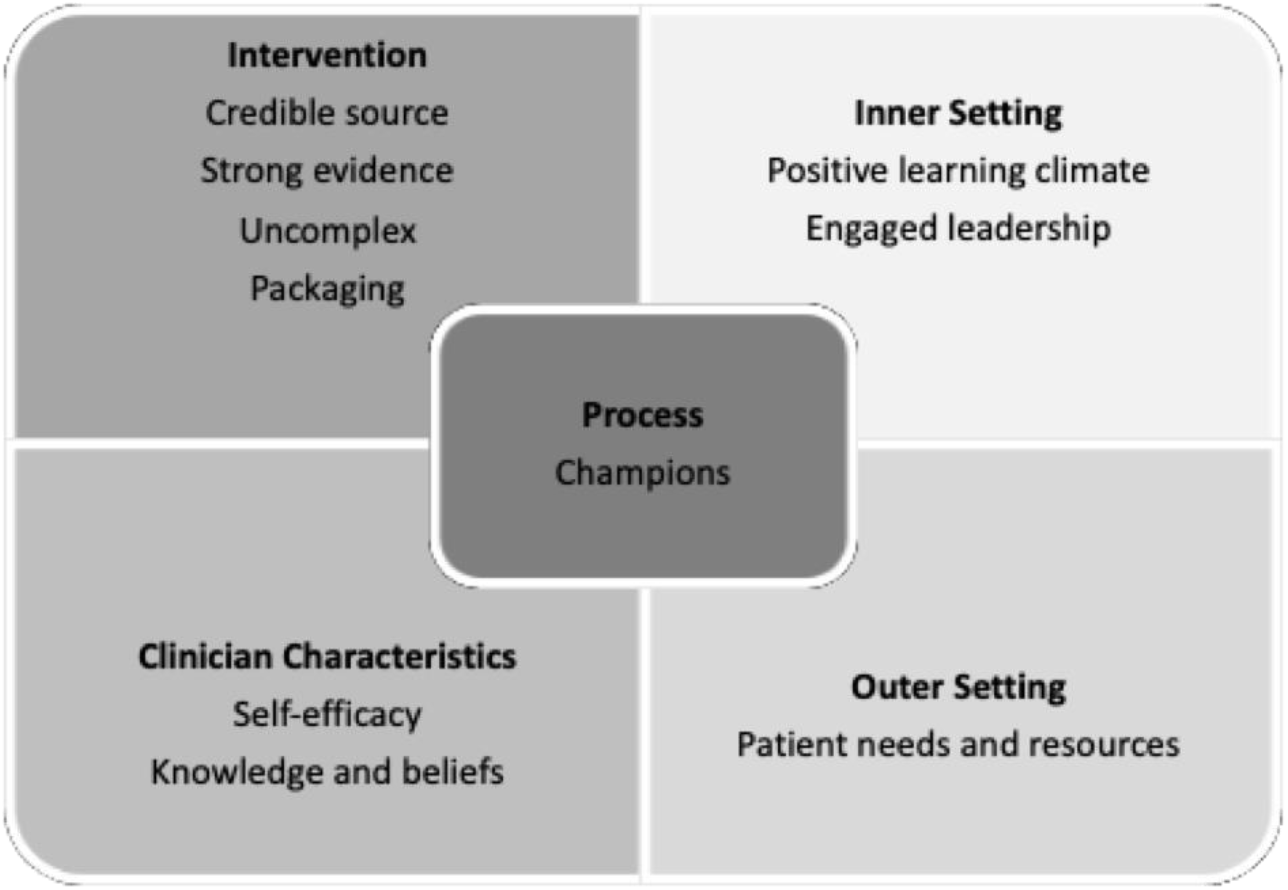
Facilitators of the PCC program as per the domains of the Consolidated Framework for Implementation Research (CFIR)

#### Intervention characteristics

Intervention characteristics of the PCC program were considered to be a strength for implementation. Specifically, these were viable intervention sources (e.g. a strong belief that the clinical supervisor had the experience necessary to provide support and feedback, and that it was clear from the beginning that the organisation had approved the intervention), quality of the evidence, the intervention was not too complex, design and packaging of the intervention (e.g. a strong agreement with the evidence base for the integrated care model). When analysed according to each component of the PCC package, clinicians clearly evaluated the workshop and supervision components much more favourably than the website and didactic seminar components.

#### Inner setting

The components of the inner setting that were a strength for implementation included fostering of a positive learning climate (e.g. the workshops provided a forum in which clinicians could pass on information to one another), along with leadership engagement. Organisational incentives and rewards appeared to have a negative impact on the implementation process.

#### Outer setting factors

These factors were a mild strength of the implementation, especially with regards to the consideration given to patient needs and resources.

#### Characteristics of individuals

The characteristics of the clinicians such as knowledge and beliefs about the intervention, self-efficacy, and ‘other personal attributes’ revealed mixed results. Knowledge and beliefs and self-efficacy were a positive while other personal attributes (e.g. the thought that one is too senior to listen to others’ opinions) negative.

#### Implementation process

components of the implementation process that were important and effective included the inclusion of clinical champions.

## DISCUSSION

This study aimed to i) evaluate the implementation of the PCC training package to improve clinician clinician practice (identification and treatment), confidence (self-efficacy), knowledge and attitudes with regards to comorbid substance use and mental disorders; and ii) identify barriers and facilitators of the PCC program using the Consolidated Framework for Implementation Research (CFIR). The study has implications for services who manage comorbid substance use and mental disorders, a complex clinical problem often associated with poor treatment outcomes.

Findings revealed that the PCC group demonstrated a significant increase in the rate of total clinical files demonstrating identification (screening and assessment) of comorbidity, yielding a 50% increase from baseline (compared with 12% decrease in the control group) and a near significant increase in rate of treatment of comorbidity (17% increase from baseline). PCC-trained clinicians also reported significantly improved levels of self-efficacy relative to controls before and after the implementation period, whereby psychologists were more likely to demonstrate these improvements. Significant improvements in knowledge and attitudes regarding screening and monitoring of symptoms associated with mental disorders were also observed in PCC-trained clinicians versus control which mirrors the clinical practice improvements with regards to identification. Exploratory analyses revealed that the improvement in clinician confidence to manage comorbidity occurred following Phase 2 during the telephone supervision and clinical champion workshops, given there was no significant improvement following Phase 1 and no additional improvement following Phase 3 booster sessions. This suggests that improvements in self-efficacy were most marked following supervision and clinical champion workshops (interactive training) relative to the website and seminars (didactic training).

There has been limited research previously conducted to improve the management of mental disorders in AOD settings. The observed improvements in clinician practice of screening and assessment in the current study resemble one previous study by Lee et al ^21^ conducted in an AOD setting. This study, while not examining implementation outcomes, evaluated the effectiveness of training for screening and brief intervention for comorbid substance use and mental disorders. These authors demonstrated significant improvements in identification, case formulation and treatment plans following a two-day workshop and clinical supervision ^21^. The observed improvements in self-efficacy in the current results are also consistent with a previous study by Hughes et al ^22^ which conducted a brief training course and follow-up supervision for case managers of community mental health teams ^22^. These authors found significant improvements in self-efficacy but not regarding changes in attitudes and did not systematically evaluate facilitators or barriers of implementation.

Our observation that improvements in self-efficacy may be specifically associated with the implementation of ongoing workshops and supervision, rather than seminars and online materials, is supported by the Motivational Interviewing (MI) training literature in this population. In particular, it is consistent with Miller and Mount’s (2011) ^40^ conclusion that a ‘one-shot’ training workshop is unlikely to alter practice behaviour significantly, along with the suggestion that workshops may not be sufficient to produce large improvements in clinician behaviour without some form of guided practice and supervision over time ^41,42^. Similar conclusions have been drawn from a study of mental health clinicians receiving comorbidity training involving cognitive behavioural therapy (CBT) workshops and a clinical champion, which were sustained over time ^43^, and in a CBT intervention for drug and alcohol clinicians involving a 3-day seminar, manual and supervision (compared to manual alone); ^44^. Indeed, the multi-modal PCC package was designed accordingly, to establish a standard of knowledge with didactic material followed by clinical supervision and champions to problem solve the implementation such that observed improvements in self-efficacy following the latter training components would be expected.

The CFIR analysis revealed that the implementation of the PCC package was mainly facilitated by strong *intervention characteristics* (credible source, uncomplicated approach, high quality design and convincing evidence), *outer setting* (good consideration of patient needs and resources), and *inner setting* (creation of a positive learning environment including allocation of time for learning and provision of appropriate and sufficient incentives, leadership engagement) factors. *Characteristics of the individuals* involved in the training had mixed effects on the implementation, as self-efficacy was a major strength, while specific personal attributes of participants weakened the impact of the implementation. The presence of clinical champions assisted with the *process* of implementation (see Figure 2).

When compared across studies of implementation barriers and facilitators of evidence-based interventions in AOD settings, the CFIR constructs identified as important in this study emulate and extend the accumulating evidence of the field. For instance, with regard to *intervention characteristics*, previous research has revealed that clinicians’ perceptions of implementation effectiveness ^45^ or a lack of clarity about the evidence behind the intervention ^46^ may influence the uptake of the intervention, and that complex guidelines can be inhibitive ^47^. This study extends these findings by suggesting that having a credible source of information is just as important as having convincing evidence for it, and by demonstrating the importance of uncomplicated psychotherapeutic approaches that are presented in a palatable format. It is possible that the addition of two interventions for separate disorders together can create complexity and uncertainty for the clinician so practical supervision regarding how to prioritise and integrate the content of treatment may be important ^48^.

Findings related to the *outer* and *inner setting* in this study corroborate previous findings in the AOD implementation literature about the importance clinicians place on patient needs and preferences when deciding whether or not to implement what might be considered to be a new intervention ^46,49,50^, along with findings about the importance of strong organisational learning climates that involve supportive training and supervision from directors and supervisors such as allocated time for learning ^49,51-53^.

Previous studies evaluating *characteristics of individuals* have identified barriers such as a lack of knowledge about evidenced-based approaches ^47^ or facilitators such as having more formal training ^54,55^, positive attitudes towards ^56,57^ or increased exposure to ^58^ evidence-based treatments, and an increased willingness to try new practices ^49^. In contrast, findings from the current study suggest that self-efficacy can be a powerful facilitator of the implementation. Another important departure from existing research is the finding that specific personal attributes of the individuals involved (such as feeling under-valued, feeling vulnerable or having a particular practice habit), may present barriers to implementation and warrant further investigation.

Lastly, while there is limited existing research into the *process* domain ^59^, clinical champions have generally been perceived as a facilitator of implementation efforts ^60-63^. There is also evidence to suggest that clinical champions contribute to a faster uptake and sustained use of the intervention ^64^, and that they can assist with generating enthusiasm amongst staff, despite systemic barriers ^65-67^.

### Limitations

Limitations include the small sample size and the non-randomised design which could not adequately account for baseline differences between PCC and control. Nonetheless, where possible we examined change from baseline and it is unlikely that other factors could be attributed to the change. In addition, the use of clinical files as a means of measuring practice change in terms of gaining an accurate insight into the actual practices of clinicians may be limited. For example, in cases where there was a large degree of variability with regards to the quality and quantity of clinical notes, there may be floor effects such that measurement of change in practice, particularly treatment, be difficult to determine accurately. Clustering if not taken into account in analyses

## Conclusion

The PCC training package was an effective means of improving the rate of comorbidity identification, increasing clinician self-efficacy regarding managing comorbidity, and influencing attitudes towards screening and assessment of comorbidity.

### Implications for Behavioral health

We evaluated the impact of the Pathways to Comorbidity Care (PCC) training program for alcohol and other drug (AOD) clinicians to improve the management of comorbid substance use disorders. Our results revealed that the implementation of the PCC package was facilitated by provision of a credible clinical supervisor, strong leadership engagement and an active clinical champion. In accordance with the barriers identified, future comorbidity training programs should consider consultation of management and peers to identify personal attributes of clinicians that may be resistant to training, and to ensure a positive learning environment within the organisation such as allocated time for learning.

## Data Availability

The datasets used and/or analysed during the current study are available from the corresponding author on reasonable request

## Acknowledgements

The authors report no acknowledgements

